# Prospective validation and comparison of clinical prediction models for early trauma care: A multicentre cohort study

**DOI:** 10.64898/2026.02.27.26347303

**Authors:** Aroke Anna Anthony, Kelvin Szolnoky, Johanna Berg, Girish Bakhshi, Debojit Basak, Nitin Borle, Shamita Chatterjee, Surbhi Chauhan, Monty Khajanchi, Tamal Khan, Anurag Mishra, L N Mohan, Sanjay Nagral, Nobhojit Roy, Rajdeep Singh, Martin Gerdin Wärnberg

**Affiliations:** World Health Organization Collaborating Centre for Emergency, Critical and Operative Care, Program for Global Surgery & Trauma, The George Institute for Global Health, New Delhi, India; Department of Medical Epidemiology and Biostatistics, Karolinska Institutet, Stockholm, Sweden; Department of Global Public Health, Karolinska Institutet, Stockholm, Sweden; Emergency Medicine, Department of Internal and Emergency Medicine, Skåne University Hospital, Malmö, Sweden; Department of General Surgery, Grant Medical College and Sir Jamshedjee Jeejeebhoy Group of Hospitals, Mumbai, India; The George Institute for Global Health, New Delhi, India; Department of Surgery, Topiwala National Medical College, Mumbai, India; Department of General Surgery, Institute of Post Graduate Medical Education and Research, Kolkata, India; Torrent Pharmaceuticals Pvt Ltd, Ahmedabad, India; Department of Surgery, Seth G. S. Medical College and K.E.M. Hospital, Mumbai, India; All India Institute of Medical Sciences, Patna, India; Department of General Surgery, Maulana Azad Medical College, New Delhi, India; Vydehi Institute of Medical Sciences and Research Centre, Bangalore, India; Department of Surgical Gastroenterology, Jaslok Hospital & Research Centre, Mumbai, India; Perioperative Medicine and Intensive Care, Karolinska University Hospital, Solna, Sweden

## Abstract

**Objective:** We aimed to prospectively validate and compare published prediction models and clinician-assigned triage categories for early trauma care.

**Design:** Prospective multicentre cohort study.

**Setting:** Three public hospitals in urban India: one secondary care hospital in Mumbai and one tertiary care teaching hospitals in Delhi and Kolkata each.

**Participants:** Adult patients aged over 18 years presenting to the emergency department with a history of trauma between 2016 and 2022. A total of 13,041 patients were included in the final analysis.

**Methods:** We externally validated five published trauma prediction models (GAP, Gerdin, KTS, MGAP, and RTS) and clinician-assigned triage categories based on initial assessment. The primary outcome was 30-day all-cause mortality. Models were recalibrated using a separate updating sample prior to evaluation, and model performance was assessed in terms of discrimination (AUC), calibration (calibration slope and plots), and decision curve analysis.

**Results:** All models and clinician gestalt-based triage demonstrated excellent discrimination (AUC range: 0.90-0.96) and good calibration after updating. The GAP model achieved the highest AUC (0.96, 95% 0.94-0.97), and RTS demonstrated the highest sensitivity (0.70).

**Conclusion:** Simple, physiology-based prediction models and clinician gestalt both demonstrated excellent performance in predicting 30-day mortality among adult trauma patients in Indian emergency departments. These findings provide a practical foundation for further development of trauma triage systems.

## Introduction

Trauma, the clinical entity composed of physical injury and the body’s physiological response^1^, is a major contributor to global mortality and morbidity. It accounts for approximately 4.4 million deaths annually, or 8% of all deaths worldwide^2^. Most of these deaths occur in low- and middle-income countries, where trauma systems are often underdeveloped and prehospital care is limited^3–5^. A key challenge in early trauma care is triage, the process of prioritising patients so that those in most urgent need are attended to first^6–8^. Without reliable triage protocols, care may be delayed and outcomes worsened for critically injured patients.

Prediction models can support triage decisions and form the basis of national trauma triage protocols in several high-income countries. These models use statistical methods to estimate the risk of outcomes such as mortality, based on clinical and demographic variables. Several models have been developed for use both prehospitally and in emergency departments. Tools such as the Revised Trauma Score (RTS), Kampala Trauma Score (KTS), MGAP, and the simplified Malawi Trauma Score have been developed or adapted for use in LMICs^10–14^. These models aim to improve feasibility by relying on a small number of easily obtainable variables.

Few studies have prospectively and independently validated these models using real-world data^15–18^. Systematic reviews have identified further limitations in existing trauma prediction models, including poor external validation, inadequate handling of missing data, and limited derivation across diverse trauma populations^10,11^. We aimed to prospectively validate and compare published prediction models and clinician-assigned triage categories for early trauma care. Our goal was to identify which models may most accurately support decision-making in early trauma care.

## Methods

### Study Design

We conducted a prospective, multicentre cohort study to validate and compare the performance of six previously published trauma prediction models: GAP^19^, Gerdin^13^, KTS^20^, MGAP^21^, Perel^12^, and RTS^22^. We also evaluated clinician-assigned triage categories. A summary of the models is presented in detail in Table 1.

**Table 1.** Models validated and compared.

| Model | Target setting | Target population | Predictors | Outcome |
| --- | --- | --- | --- | --- |
| GAP<sup>19</sup> | Hospital | All trauma patients | GCS, cAge, cSBP | In-hospital mortality |
| Gerdin et al.<sup>13</sup> | Hospital | Trauma, aged > 14 years | SBP, GCS, HR | In-hospital 24h mortality |
| KTS<sup>20</sup> | Hospital | All trauma patients | cSBP, cRR, LOC, Number of serious injuries | Composite of mortality and need for admission |
| MGAP<sup>21</sup> | Prehospital | All trauma patients | GCS, cSBP, MOI, cAge | In-hospital mortality |
| Perel et al.<sup>12</sup> | Hospital | Patients with traumatic bleeding | GCS, Age, SBP, TXA | In-hospital mortality within 28 days after injury |
| T-RTS<sup>22</sup> | Prehospital | All trauma patients aged > 14 years | cSBP, cRR, cGCS | In-hospital mortality |

**Note** c: Categorised; GAP: Glasgow Coma Scale, Age and Systolic Blood Pressure; GCS: Glasgow Coma Scale; HR: Heart Rate; KTS: Kampala Trauma Score; LOC: Level of Consciousness; MGAP: Mechanism, Glasgow Coma Scale, Age and Systolic Blood Pressure; MOI: Mechanism of Injury; RR: Respiratory Rate; RTS: Revised Trauma Score; SBP: Systolic Blood Pressure.

### Setting

Data were collected between July 2016 and April 2022 from three public hospitals in urban India: Khershedji Behramji Bhabha Hospital (KBBH), a secondary care hospital in Mumbai; Lok Nayak Hospital of Maulana Azad Medical College (MAMC) in Delhi; and the Institute of Post-Graduate Medical Education and Research and Seth Sukhlal Karnani Memorial Hospital (SSKM) in Kolkata, both tertiary care teaching hospitals.

### Participants

#### Eligibility Criteria

We included patients aged 18 & older presenting to the emergency department (ED) with a history of trauma, defined using the International Classification of Diseases version 10 (ICD-10), Chapter 20 codes V01–Y36. Patients with external causes not meeting the definition of trauma, such as iatrogenic injuries or poisonings, were excluded.

#### Sampling and Follow-Up

At each site, trained research officers prospectively enrolled the first ten eligible patients presenting during the first six hours of their shift. It was decided to include only the first ten patients to ensure that data collection, including follow up, was feasible. Shifts rotated between morning, evening, and night, and the order of rotation was randomized. Research officers worked five shifts per week. Follow-up assessments were conducted at 24 hours, 30 days, and 6 months after ED arrival, using hospital records and structured telephone interviews with patients or their relatives.

### Outcomes

The outcome was 30-day all-cause mortality.

### Predictors

Predictor variables were defined based on the specifications of the included models and included systolic blood pressure (SBP), heart rate (HR), respiratory rate (RR), Glasgow Coma Scale (GCS), AVPU score, age, number of serious injuries, and mechanism of injury. Variables were recorded at ED arrival by research officers using standardised case record forms and calibrated instruments. Where possible, definitions followed the Utstein Trauma Template^23^. To capture clinician gestalt, treating clinicians were asked to assign an initial triage category to each patient at the time of presentation. Triage categories were based on clinical judgement of acuity and recorded as one of four colour codes (green, yellow, orange, or red), with green indicating the least critical cases and red the most critical. No formal triage scoring tool was used.

### Handling of Missing Data

It was initially planned to use multiple imputation using chained equations to handle missing data, but because of very low missing data frequencies in predictor variables, we decided to conduct a complete case analysis instead. We compared the baseline characteristics between patients with complete and incomplete data to assess potential bias due to missing data. For continuous variables, Welch’s two-sample t-test was used to compare group means, and standardised mean differences (SMDs) were calculated to assess the magnitude of differences. For categorical variables, two-sample tests for equality of proportions with continuity correction were performed.

### Statistical Analysis

The study samples are described using frequencies and percentages for qualitative variables and median and interquartile ranges for quantitative variables. Model performance was assessed in two stages - model updating, and model validation. The data was first split into an updating sample, used to recalibrate the models, and a validation sample, used to test how well the updated models perform. Each model was recalibrated using logistic regression in the updating sample, consisting of the first 75 outcome events (patients with outcome) and a proportionate number of non-events presenting during the same time period to preserve the observed event rate. We based the size of this updating sample on our previous work^24^. The recalibrated models were evaluated in the remaining validation sample. Model performance was assessed in terms of discrimination, calibration, clinical utility and classification accuracy.

Discrimination was assessed using the area under the receiver operating characteristic curve (AUC). We compared predicted vs. observed probabilities across deciles of risk to assess calibration. We generated calibration plots and estimated calibration slopes using logistic recalibration. Clinical utility was assessed using decision curve analysis^25^ and classification accuracy, based on sensitivity and specificity at a threshold of 0.5, and mistriage rates.

All models were compared in a pairwise fashion. We computed 95% confidence intervals from 1000 bootstrap resamples. All analyses were performed in R version 4.5.0 (2025-04-11).

### Quantitative Variables

Continuous predictors were analysed in their original form unless categorisation was required for score calculation, in which case categories followed the original model specifications. Where appropriate, non-linear associations were modelled using restricted cubic splines, consistent with the development of the original models.

### Quality control measures

We implemented several quality control measures. Weekly virtual meetings were held between the project management team and site-based research officers to review recently collected data and resolve discrepancies. Every 3–4 months, an external project officer independently collected data in parallel with a site officer for a 3-days. The two resulting datasets were compared to assess inter-rater consistency. Furthermore, case record forms had embedded validity checks, and all measurement equipment was routinely calibrated.

### Sample Size

The study protocol included outcome assessments at 24 hours and 6 months, which will be reported separately, and the sample size calculations were informed by the rarest outcome of interest, 24-hour mortality, anticipated to occur in 5% of patients. Each centre aimed to enroll 4000 patients to ensure 200 events, allowing for model comparison with 80% power. After adjusting for up to 20% loss to follow-up, the target sample size was 5400 patients per centre. Given an enrolment rate of 10 patients per shift and 5 shifts per week, the estimated duration for accrual was 27 months. For model updating, a sample of 75 events and corresponding non-events was used, based on prior work showing this to be sufficient for recalibrating logistic models with up to eight parameters^24,26^.

### Ethical Considerations

Ethical approval was obtained from the relevant institutional review boards. As the study was observational and involved no intervention, informed consent was waived for initial data collection involving vital signs and demographic parameters. Informed consent was obtained for follow-up. For patients unable to provide consent at discharge (e.g., due to GCS < 15), consent was obtained from a relative or legally authorised representative. The research was approved by the ethics review board at each participating hospital. The names of the boards and the approval numbers are as follows: the Ethics and Scientific Committee (KBBH, HO/4982/KBB), the Institutional Ethics Committee (MAMC, F.1/IEC/MAMC/53/2/2016/No97) and the IPGME&R Research Oversight Committee (SSKM, Inst/IEC/2016/328). The data management and analysis were also approved by the Stockholm Regional Ethics Committee (2018/2201-31/2).

## Results

### Participants

Of 15,366 eligible patients, 13,041 patients had complete data and were included in the final analysis. The extent of incomplete cases for each variable and a comparison of complete and incomplete data is reported in Appendix 1, showing that the two groups were largely similar in demographics and physiological characters. The mean age was 32 years (SD 24, 45), and 9,978 patients (77%) were male. The majority of injuries were caused by blunt trauma, and most patients were alert on the AVPU scale. Serious injuries, as determined by the treating physician according to KTS, were present in a minority of patients, with 61% having no serious injuries and 5.5% having multiple. Mortality within 30 days was observed in 4.7% of the cohort, while in-hospital mortality occurred in 2.9%, 24-hour mortality in 1.9%, and 6-month mortality in 6%. Hospital admission was recorded for 9.3% of patients, and ICU admission for 0.4% (Table 1). Comparing patients that were alive or dead at 30 days, median GCS was notably lower in the group that died, 7 vs 15, suggesting that GCS is an important predictor of mortality in this cohort (Table 1).

**Table 2:** Sample characteristics, stratified by 30-day all-cause mortality.

| Characteristic | Alive N = 12,428<sup>1</sup> | Dead N = 613<sup>1</sup> | Overall N = 13,041<sup>1</sup> |
| --- | --- | --- | --- |
| Age | 32 (24, 45) | 45 (28, 60) | 32 (24, 45) |
| Sex |  |  |  |
| Female | 2,938 (24%) | 122 (20%) | 3,060 (23%) |
| Male | 9,488 (76%) | 490 (80%) | 9,978 (77%) |
| Unknown | 2 | 1 | 3 |
| Glasgow coma scale | 15.00 (15.00, 15.00) | 7.00 (4.00, 9.00) | 15.00 (15.00, 15.00) |
| Systolic blood pressure | 124 (116, 132) | 111 (99, 129) | 124 (115, 132) |
| Heart rate | 82 (76, 92) | 89 (77, 109) | 82 (76, 92) |
| Respiratory rate | 21.0 (19.0, 24.0) | 20.0 (16.0, 22.0) | 21.0 (19.0, 23.0) |
| Type of injury |  |  |  |
| Blunt | 12,277 (99%) | 606 (99%) | 12,883 (99%) |
| Penetrating | 144 (1.2%) | 5 (0.8%) | 149 (1.1%) |
| Both blunt and penetrating | 7 (<0.1%) | 2 (0.3%) | 9 (<0.1%) |
| Number of serious injuries |  |  |  |
| Nil/no serious injury | 7,931 (64%) | 9 (1.5%) | 7,940 (61%) |
| Single serious injury | 3,948 (32%) | 430 (70%) | 4,378 (34%) |
| Multiple serious injuries | 549 (4.4%) | 174 (28%) | 723 (5.5%) |
| AVPU |  |  |  |
| Unresponsive | 9 (<0.1%) | 142 (23%) | 151 (1.2%) |
| Pain responsive | 286 (2.3%) | 383 (62%) | 669 (5.1%) |
| Voice responsive | 187 (1.5%) | 29 (4.7%) | 216 (1.7%) |
| Alert | 11,946 (96%) | 59 (9.6%) | 12,005 (92%) |
| Dead at 24 hours | 0 (0%) | 249 (41%) | 249 (1.9%) |
| Unknown | 0 | 1 | 1 |
| Dead at 90 days | 42 (0.4%) | 613 (100%) | 655 (6.0%) |
| Unknown | 2,167 | 0 | 2,167 |
| In-hospital mortality | 0 (0%) | 377 (62%) | 377 (2.9%) |
| Unknown | 58 | 2 | 60 |
| Admitted to the hospital | 826 (6.7%) | 392 (64%) | 1,218 (9.3%) |
| Unknown | 9 | 0 | 9 |
| Admitted to the intensive care unit | 17 (0.1%) | 34 (5.6%) | 51 (0.4%) |
| Unknown | 11 | 4 | 15 |
| Clinician-assigned triage category |  |  |  |
| Green | 7,141 (58%) | 7 (1.2%) | 7,148 (55%) |
| Yellow | 4,172 (34%) | 138 (23%) | 4,310 (33%) |
| Orange | 777 (6.3%) | 191 (32%) | 968 (7.5%) |
| Red | 267 (2.2%) | 259 (44%) | 526 (4.1%) |
| Unknown | 71 | 18 | 89 |

<sup>1</sup>Median (Q1, Q3); n (%)

### Discrimination, Calibration, and Clinical Utility

The predictive performance of each recalibrated model for 30-day mortality is shown in Table 3. Discrimination, as measured by the AUC, across models ranged from 0.90 (Perel) to 0.96 (GAP). AUC was highest for the GAP model (AUC 0.96), followed closely by MGAP (AUC 0.95), KTS (AUC 0.95), and RTS (AUC 0.95) (Figure 1). The Gerdin model also performed well (AUC 0.94) while the Perel and clinician-assigned triage categories showed comparatively lower discrimination (AUCs 0.90 and 0.91). Calibration slopes ranged from 0.77 (Perel) to 1.07 (MGAP, GAP), and were close to one for KTS, RTS and Gerdin models.

**Figure 1:**
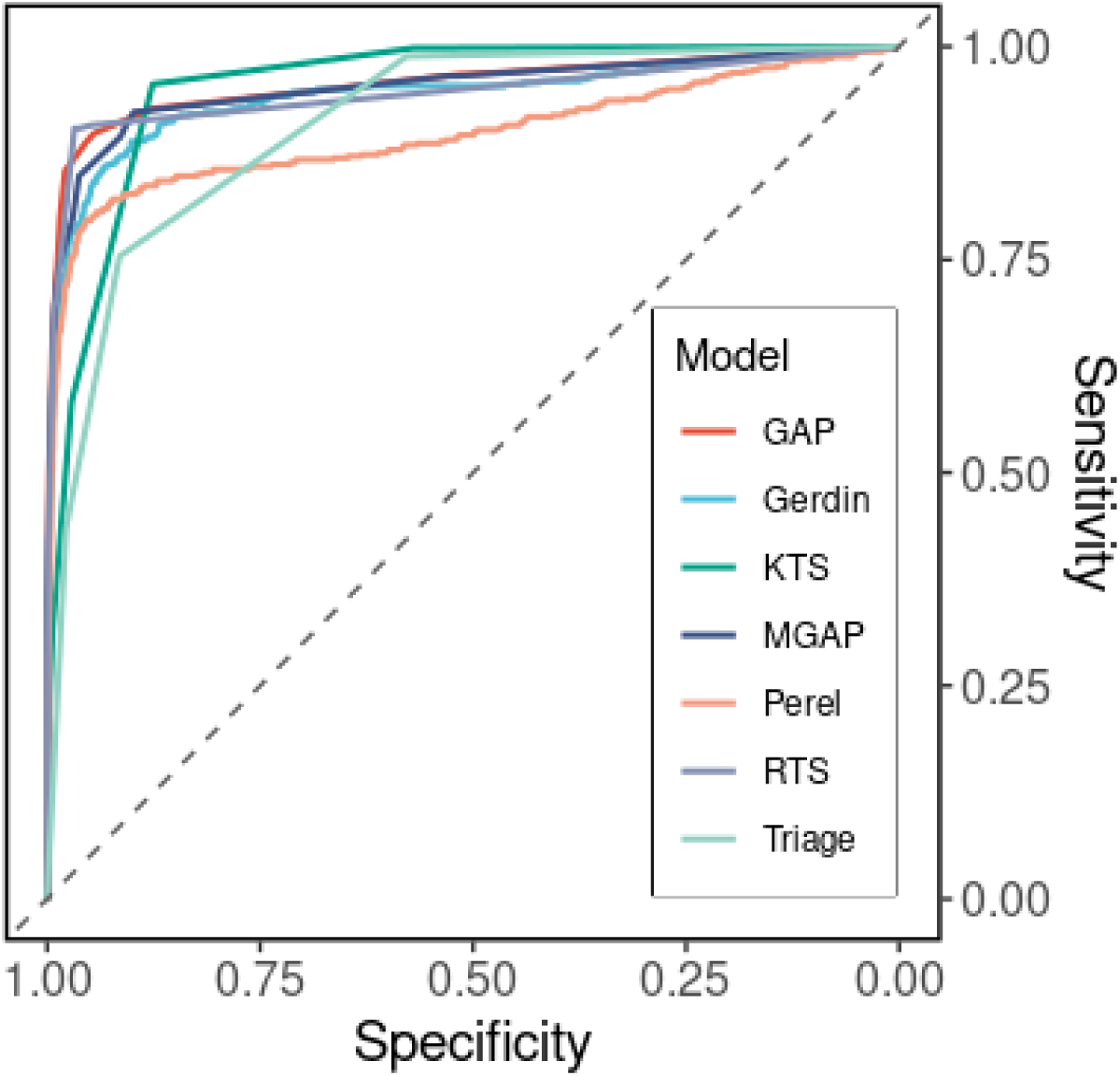
Receiver operating characteristics curves for all models. Triage refers to the clinician-assigned triage categories. Abbreviations: GAP: Glasgow Coma Scale, Age and Systolic Blood Pressure; KTS: Kampala Trauma Score; MGAP: Mechanism, Glasgow Coma Scale, Age and Systolic Blood Pressure; RTS: Revised Trauma Score.

**Table 3:** Summary performance and predictors by model.

| Model | AUC (95% CI) | Sensitivity | Specificity | Calibration Slope | SBP | HR | RR | GCS | AVPU | Age | Penetrating | NSI |
| --- | --- | --- | --- | --- | --- | --- | --- | --- | --- | --- | --- | --- |
| **GAP** | 0.96 (0.94–0.97) | 0.67 | 0.99 | 1.07 (1.01–1.13) | ✓ | - | - | ✓ | - | ✓ | - | - |
| **Gerdin** | 0.94 (0.93–0.96) | 0.54 | 1.00 | 0.97 (0.90–1.04) | ✓ | ✓ | - | ✓ | - | - | - | - |
| **KTS** | 0.95 (0.95–0.96) | 0.31 | 0.99 | 1.01 (0.95–1.08) | ✓ | - | ✓ | - | ✓ | ✓ | - | ✓ |
| **MGAP** | 0.95 (0.94–0.96) | 0.60 | 0.99 | 1.07 (1.00–1.13) | ✓ | f- | - | ✓ | - | ✓ | ✓ | - |
| **Perel<sup>1</sup>** | 0.90 (0.87–0.91) | 0.50 | 0.99 | 0.77 (0.71–0.82) | ✓ | ✓ | ✓ | ✓ | - | ✓ | ✓ | - |
| **RTS** | 0.95 (0.93–0.96) | 0.70 | 0.99 | 0.97 (0.91–1.03) | ✓ | - | ✓ | ✓ | - | - | - | - |
| **Triage<sup>2</sup>** | 0.91 (0.90–0.92) | 0.43 | 0.98 | 0.94 (0.88–1.00) | - | - | - | - | - | - | - | - |

<sup>1</sup>We implemented the simplified Perel-model, and had no information on TXA use
<sup>2</sup>Clinician-assigned triage category (Red, Orange, Yellow, Green).
Abbreviations: GAP: Glasgow Coma Scale, Age and Systolic Blood Pressure; KTS: Kampala Trauma Score; MGAP: Mechanism, Glasgow Coma Scale, Age and Systolic Blood Pressure; RTS: Revised Trauma Score

Visual inspection of calibration plots available in Appendix 2 revealed that the Perel model and clinician-assigned triage categories slightly underestimated risk at higher predicted probabilities (Slopes of 0.77 and 0.94). The GAP and MGAP calibration slopes were slightly above 1, indicating mild over-prediction. RTS demonstrated the highest sensitivity (0.70), and specificity ranged from 0.98 to 1.00. Pairwise comparisons of AUC values are shown in Figure 2. While point estimates differed slightly between models, most confidence intervals overlapped. GAP and RTS consistently outperformed other models by small but statistically significant margins.

**Figure 2:**
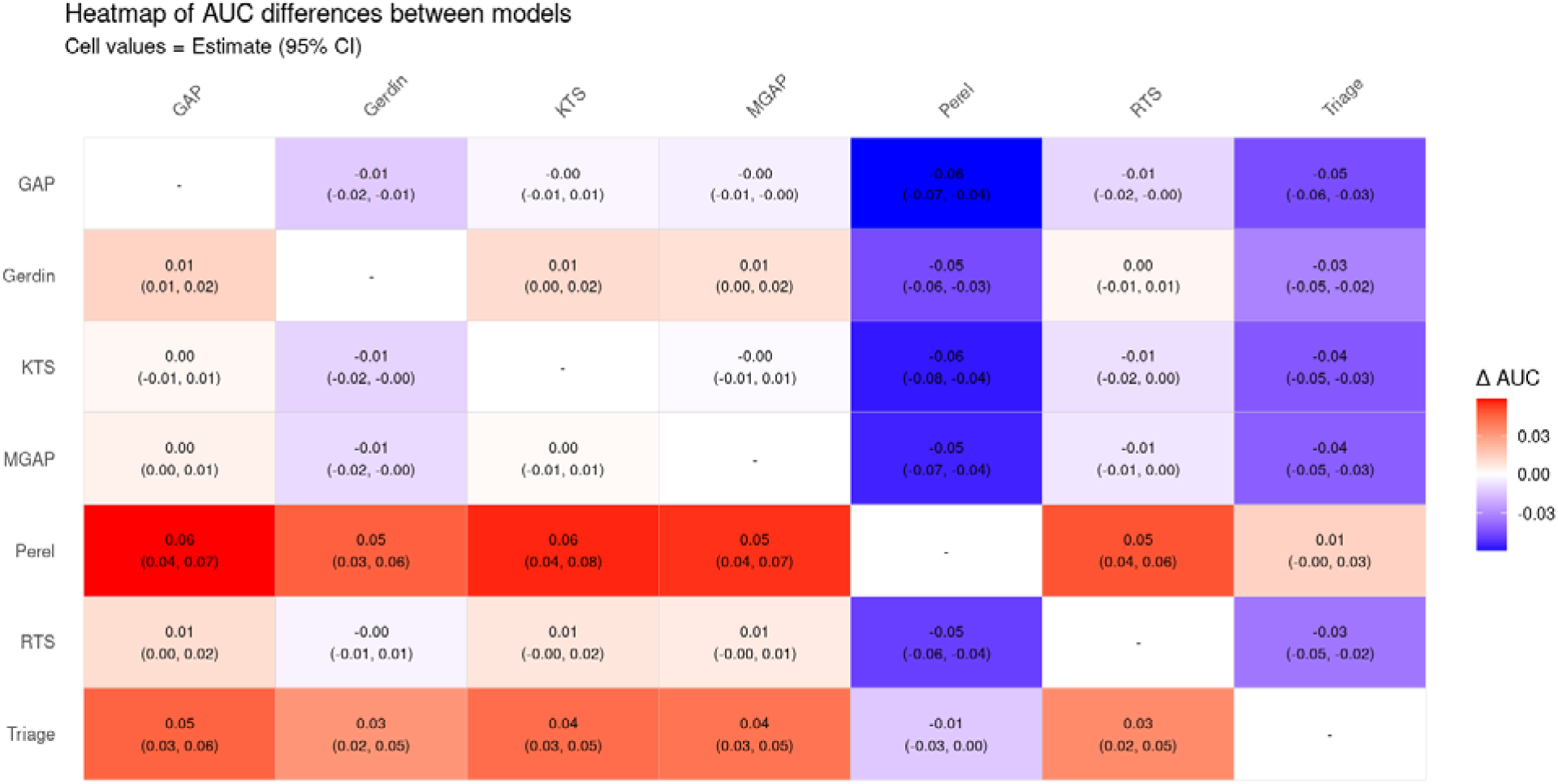
Heatmap of differences in area under the receiver operating characteristic curve (AUC) between models. Triage refers to the clinician-assigned triage categories. Red color indicates positive differences and blue color indicates negative differences. For example, the red cell in the GAP column on the Perel row shows that the GAP model’s AUC was 0.06 points higher than the Perel model’s AUC, and that the 95% confidence interval (CI) was 0.04 to 0.07 points, pointing to a small but statistically significant difference. Abbreviations: GAP: Glasgow Coma Scale, Age and Systolic Blood Pressure; KTS: Kampala Trauma Score; MGAP: Mechanism, Glasgow Coma Scale, Age and Systolic Blood Pressure; RTS: Revised Trauma Score.

Figure 3 presents a decision curve analysis comparing the net benefit of the models across a range of threshold probabilities. Across most thresholds, models such as MGAP, GAP and RTS demonstrate better net benefit compared to the default strategies of treating all or treating none. MGAP and GAP demonstrated particularly strong performance at moderate threshold probabilities (30-60%). Clinician-assigned triage categories and models such as Perel and KTS exhibited lower net benefit across most threshold probabilities.

**Figure 3:**
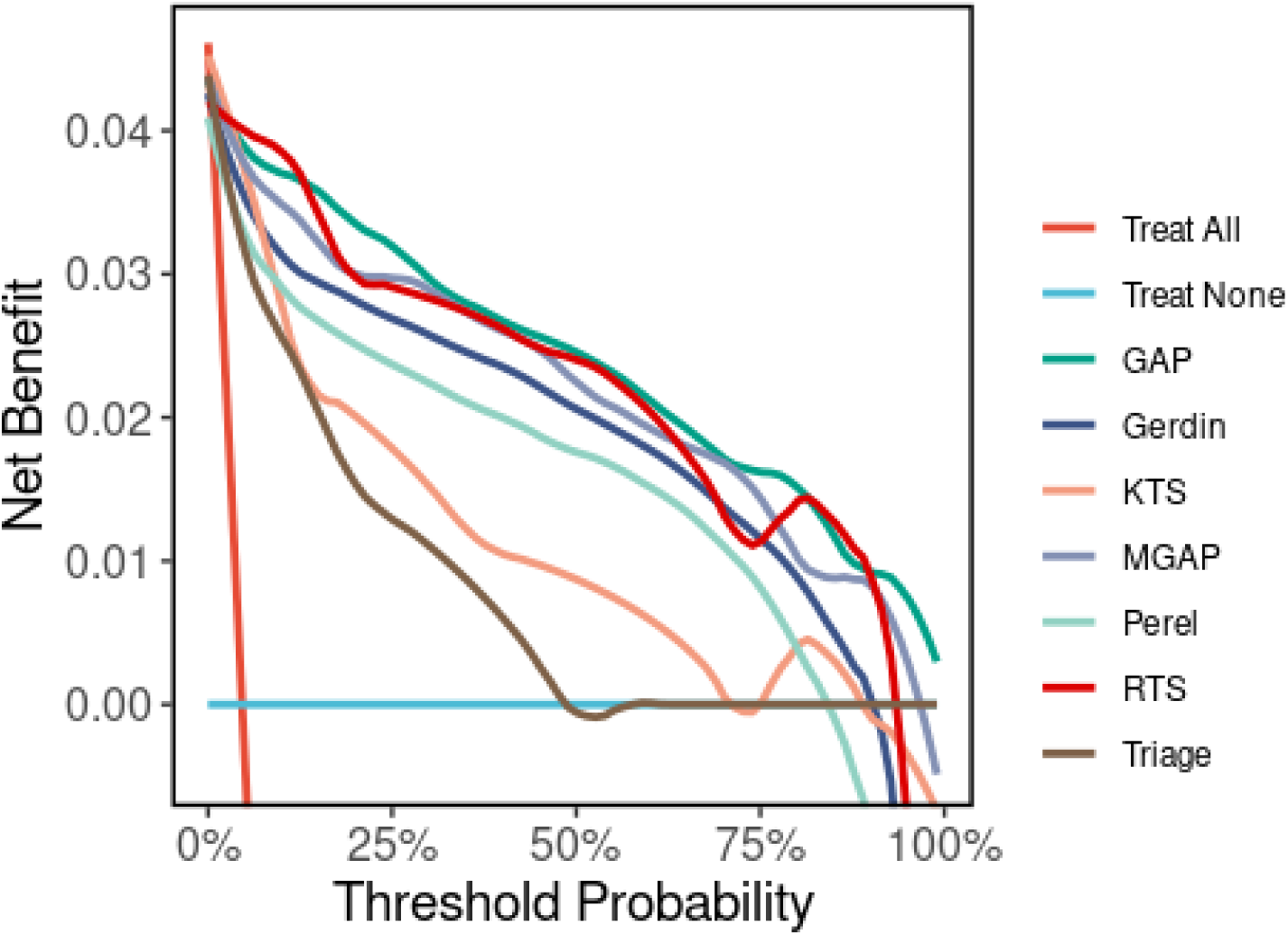
Decision curve analysis. Triage refers to the clinician-assigned triage categories. Abbreviations: GAP: Glasgow Coma Scale, Age and Systolic Blood Pressure; KTS: Kampala Trauma Score; MGAP: Mechanism, Glasgow Coma Scale, Age and Systolic Blood Pressure; RTS: Revised Trauma Score.

## Discussion

We found that five existing trauma prediction models, GAP, Gerdin, KTS, MGAP, and RTS as well as clinician-assigned triage categories, demonstrated excellent discrimination and good calibration when applied to adult trauma patients presenting to urban emergency departments in India. The GAP model achieved the highest AUC, and RTS the highest sensitivity, although overall differences in performance were modest. Notably, models relying on a small number of physiological variables, such as systolic blood pressure, GCS, and age, were sufficient to produce reliable estimates of 30-day mortality risk. It is important to note that the clinician-assigned triage categories reflected perceived clinical urgency rather than an explicit prediction of mortality. Clinicians were instructed to assign a triage colour according to urgency of care, not risk of death. As such, comparisons between model-predicted mortality and clinician judgement should be interpreted with caution. Some patients assigned a high-priority triage category may have survived because they were identified and treated early, resulting in lower observed mortality than their initial acuity would suggest. In this context, superior discrimination of mortality risk by the prediction models does not necessarily imply better real-time triage performance. In a separate study using an earlier subset of this cohort, we compared clinician gestalt with prediction models using a fixed cutoff and found no significant difference in discrimination between clinical judgement and physiology-based scores^27^.

Our findings are consistent with previous validations of simplified trauma scores in LMIC settings, which have shown that physiology-based models perform well even when resources and data quality are limited.^28^ In our setting, the best-performing models used variables readily available at the bedside, requiring no advanced imaging or laboratory results. This makes them particularly well-suited for integration into routine practice in emergency departments. Many hospitals in urban India serve large populations and receive both direct and referred trauma cases, often without a structured triage system. In such settings, adopting a simple and validated scoring model could support more consistent triage decisions, help identify high-risk patients earlier, and improve time to critical interventions. Evidence on clinician gestalt for triage is limited and mixed. An U.S. study suggests limited sensitivity of gestalt for ruling out injury. In comparison, a cross-context model validation showed that algorithmic models detain discrimination but require re-calibration for new setting.^24^ Studies emphasise that clinical judgement can guide triage^28,29^ but should be supported by structured decision aids that improve accuracy and reduce variability^30^. These findings highlight the potential role of prediction models as decision support tools to complement, rather than replace, clinical assessment in resource-constrained environments.

Notably, we found that all models, including those derived in high-income countries, discriminated well between patients who died and those who survived. This suggests that the physiological predictors of trauma mortality may be consistent across contexts, despite differences in health system capacity and patient populations. However, calibration varied somewhat, with the Perel model showing poorer performance than others, even after updating. This may reflect contextual mismatch or unmeasured variables such as the use of tranexamic acid (TXA), which was part of the original model but not collected in our dataset. This also aligns with previous research indicating that calibration is adversely affected when clinical prediction models are moved between contexts.^31^ Poor calibration could limit the clinical utility of a prediction model by mis-estimating risk levels, leading to inappropriate prioritisation of care. In the context of trauma triage, this could result in high-risk patients not being recognised in time or low-risk patients receiving unnecessary escalation of care.

An important implication of our findings is that more complex or data-intensive models are unlikely to add substantial value in LMIC trauma settings, at least when the outcome is 30 day mortality. In fact, our study supports the growing consensus that simple, interpretable models often perform as well as or better than more complex approaches, especially when implemented in real-world, resource-limited environments. It is likely that simple models generalise better because having a few, stable predictors makes them less prone to overfitting. While machine learning methods have shown promise in retrospective analyses, their benefits over traditional models in prospective LMIC cohorts remain unproven.^32–34^ They also introduce additional challenges related to implementation, transparency, and trust among clinicians.

Our study has several strengths. It is one of the first to prospectively compare multiple validated trauma models in an LMIC context using all patients presenting with trauma, not only those admitted. This enhances the generalisability of our findings and their relevance to real-world triage systems. Data collection was rigorous, with standardised measurements and extensive quality control procedures. We also used a robust updating and validation approach, ensuring that each model was recalibrated before evaluation. However, we acknowledge some limitations. First, although our sample was large and diverse, it was limited to three urban hospitals, and findings may not generalise to rural or less-resourced settings. Second, we did not collect information on TXA administration, which may have affected the calibration of the Perel model. Third, our analysis focused on 30-day mortality, but other outcomes such as disability, long-term recovery, or need for surgery may also be relevant for triage decisions. Fourth, traumatic brain injury was a major contributor to mortality in our cohort, which may have amplified the predictive strength of the GCS relative to other variables. In settings with a different trauma profile, the relative importance of predictors may vary.

Taken together, our findings provide one of the first comprehensive prospective validations of published trauma triage models in a real-world LMIC setting. We demonstrated that both simple, physiology-based models and clinician gestalt retain strong predictive performance even in resource-constrained environments. Our study offers a practical foundation for the development of formal trauma triage systems across similar contexts. In settings where structured triage protocols are lacking, these models, complemented by clinical judgement, could help strengthen emergency care delivery. Future research should explore additional outcomes, such as shorter term mortality and critical interventions, and further evaluate implementation strategies.

## Data Availability

De-identified data is available from the corresponding author on reasonable request.

## Author contributions

MGW conceived of and designed the study. AA wrote the first draft, assisted in subsequent revisions, drafted the analysis code, and conducted preliminary analyses. KS refined the analysis code and conducted the final analyses. JB further assisted with analysis. All authors provided input during the design, implementation and interpretation phases of the study. All authors provided input on the manuscript.

## Data sharing statement

De-identified data is available from the corresponding author on reasonable request.

## References

1. Gerdin M. The Risk of Dying: Predicting Trauma Mortality in Urban Indian Hospitals. Dept of Public Health Sciences; Karolinska Institutet; 2015.

2. Lou J, Xiang Z, Zhu X, et al. Trends and levels of the global, regional, and national burden of injuries from 1990 to 2021: Findings from the global burden of disease study 2021. Annals of Medicine. 2025;57(1):2537917.

3. James SL, Castle CD, Dingels ZV, et al. Global injury morbidity and mortality from 1990 to 2017: Results from the global burden of disease study 2017. Injury Prevention. 2020;26(Supp 1):i96-i114.

4. Misra S, Tiwari S. Trauma care system in India: Where are we? International Journal of Research and Analytical Reviews. 2018;5(3).

5. Babhulkar S, Apte A, Barick D, Hoogervorst P, Tian Y, Wang Y. Trauma care systems in India and China: A grim past and an evolving future. OTA International. 2019;2(S1):e017.

6. Yancey CC, O’Rourke MC. Emergency department triage. Published online 2020.

7. Granström A, Strömmer L, Schandl A, Östlund A. A criteria-directed protocol for in-hospital triage of trauma patients. European Journal of Emergency Medicine. 2018;25(1):25–31.

8. Amato S, Bonnell L, Mohan M, Roy N, Malhotra A. Comparing trauma mortality of injured patients in India and the USA: A risk-adjusted analysis. Trauma Surgery & Acute Care Open. 2021;6(1):e000719.

9. Collins GS, Reitsma JB, Altman DG, Moons KG. Transparent reporting of a multivariable prediction model for individual prognosis or diagnosis (TRIPOD) the TRIPOD statement. Circulation. 2015;131(2):211–219.

10. Rehn M, Perel P, Blackhall K, Lossius HM. Prognostic models for the early care of trauma patients: A systematic review. Scandinavian journal of trauma, resuscitation and emergency medicine. 2011;19(1):1–8.

11. Munter L de, Polinder S, Lansink KW, Cnossen MC, Steyerberg EW, Jongh MA de. Mortality prediction models in the general trauma population: A systematic review. Injury. 2017;48(2):221–229.

12. Perel P, Prieto-Merino D, Shakur H, et al. Predicting early death in patients with traumatic bleeding: Development and validation of prognostic model. Bmj. 2012;345.

13. Gerdin M, Roy N, Khajanchi M, et al. Predicting early mortality in adult trauma patients admitted to three public university hospitals in urban India: A prospective multicentre cohort study. PloS one. 2014;9(9):e105606.

14. Gallaher J, Jefferson M, Varela C, Maine R, Cairns B, Charles A. The malawi trauma score: A model for predicting trauma-associated mortality in a resource-poor setting. Injury. 2019;50(9):1552–1557.

15. Bouzat P, Legrand R, Gillois P, et al. Prediction of intra-hospital mortality after severe trauma: Which pre-hospital score is the most accurate? Injury. 2016;47(1):14–18.

16. Shao-Chun Wu SCK Cheng-Shyuan Rau. The reverse shock index multiplied by glasgow coma scale score (rSIG) and prediction of mortality outcome in adult trauma patients: A cross-sectional analysis based on registered trauma data. International journal of environmental research and public health. 2018;15(11):2346.

17. Nobhojit Roy ES Martin Gerdin. Validation of international trauma scoring systems in urban trauma centres in India. Injury. 2016;47(11):2459-2464.

18. Merchant AAH, Shaukat N, Ashraf N, et al. Which curve is better? A comparative analysis of trauma scoring systems in a south asian country. Trauma surgery & acute care open. 2023;8(1):e001171.

19. Kondo Y, Abe T, Kohshi K, Tokuda Y, Cook EF, Kukita I. Revised trauma scoring system to predict in-hospital mortality in the emergency department: Glasgow coma scale, age, and systolic blood pressure score. Critical care. 2011;15(4):1–8.

20. Kobusingye OC, Lett RR. Hospital-based trauma registries in uganda. Journal of Trauma and Acute Care Surgery. 2000;48(3):498–502.

21. Sartorius D, Le Manach Y, David JS, et al. Mechanism, glasgow coma scale, age, and arterial pressure (MGAP): A new simple prehospital triage score to predict mortality in trauma patients. Critical care medicine. 2010;38(3):831–837.

22. Champion HR, Sacco WJ, Copes WS, Gann DS, Gennarelli TA, Flanagan ME. A revision of the trauma score. The Journal of Trauma. 1989;29(5):623–629.

23. Ringdal K, Coats T, Lefering R, et al. The utstein trauma template for uniform reporting of data following major trauma: Data dictionary. 2008. European Trauma Registry Network. Published online 2008.

24. Gerdin M, Roy N, Felländer-Tsai L, et al. Traumatic transfers: Calibration is adversely affected when prediction models are transferred between trauma care contexts in India and the united states. Journal of clinical epidemiology. 2016;74:177–186.

25. Vickers AJ, Elkin EB. Decision curve analysis: A novel method for evaluating prediction models. Medical Decision Making. 2006;26(6):565–574.

26. Steyerberg EW. Clinical prediction models - a practical approach to development, validation, and updating. Published online 2016.

27. Wärnberg Gerdin L, Khajanchi M, Kumar V, et al. Comparison of emergency department trauma triage performance of clinicians and clinical prediction models: A cohort study in India. BMJ open. 2020;10(2):e032900. doi:10.1136/bmjopen-2019-032900

28. Yost MT, Carvalho MM, Mbuh L, et al. Back to the basics: Clinical assessment yields robust mortality prediction and increased feasibility in low resource settings. PLOS global public health. 2023;3(3):e0001761. doi:10.1371/journal.pgph.0001761

29. Shiber J, Fontane E, Patel J, et al. Gestalt clinical severity score (GCSS) as a predictor of patient severity of illness or injury. The American journal of emergency medicine. 2023;66:11–15. doi:10.1016/j.ajem.2023.01.005

30. Rodriguez N, Montoy JC, Mower W, et al. 61 gestalt in acute trauma: Evaluating clinician accuracy in predicting abdominal injuries among trauma patients. Annals of emergency medicine. 2021;78(4):S25. doi:10.1016/j.annemergmed.2021.09.070

31. Gerdin M, Roy N, Felländer-Tsai L, et al. Traumatic transfers: Calibration is adversely affected when prediction models are transferred between trauma care contexts in India and the united states. Journal of clinical epidemiology. 2016;74:177–186.

32. Nsubuga M, Kintu TM, Please H, Stewart K, Navarro SM. Enhancing trauma triage in low-resource settings using machine learning: A performance comparison with the kampala trauma score. BMC Emergency Medicine. 2025;25(1):14.

33. Le K, Chen J, Mai D, Le KDR. An evaluation on the potential of large language models for use in trauma triage. Emergency Care and Medicine. 2024;1(4):350–367.

34. Zhang T, Nikouline A, Lightfoot D, Nolan B. Machine learning in the prediction of trauma outcomes: A systematic review. Annals of emergency medicine. 2022;80(5):440–455.

