## Appendix for "Prospective validation and comparison of clinical prediction models for early trauma care: A multicentre cohort study"

### Appendices

Appendix 1: Table comparing patients with missing vs non-missing mortality

| **Characteristic** | **Complete** N = 13,356*^1^* | **Incomplete** N = 2,010*^1^* | **Difference***^2^* | **95% CI***^2^* | **p-value***^2^* |
| --- | --- | --- | --- | --- | --- |
| Age | 32 (24, 45) | 32 (24, 45) | 0.50 | -0.17, 1.2 | 0.14 |
| Sex |  |  | 0.04 | 0.00, 0.09 |  |
| Female | 3,123 (23%) | 433 (22%) |  |  |  |
| Male | 10,230 (77%) | 1,577 (78%) |  |  |  |
| Glasgow coma scale | 15.00 (15.00, 15.00) | 15.00 (15.00, 15.00) | -0.36 | -0.43, -0.30 | <0.001 |
| Systolic blood pressure | 124 (114, 132) | 122 (112, 131) | 0.81 | 0.01, 1.6 | 0.047 |
| Heart rate | 82 (76, 92) | 84 (78, 95) | -1.6 | -2.3, -0.89 | <0.001 |
| Repiratory rate | 21.0 (19.0, 24.0) | 21.0 (19.0, 23.0) | -0.05 | -0.20, 0.10 | 0.5 |
| Type of injury |  |  | 0.05 | 0.00, 0.09 |  |
| Blunt | 13,193 (99%) | 1,972 (98%) |  |  |  |
| Penetrating | 153 (1.1%) | 34 (1.7%) |  |  |  |
| Both blunt and penetrating | 10 (<0.1%) | 2 (<0.1%) |  |  |  |
| Number of serious injuries |  |  | 0.31 | 0.27, 0.36 |  |
| Nil/no serious injury | 7,969 (61%) | 1,490 (75%) |  |  |  |
| Single serious injury | 4,436 (34%) | 426 (21%) |  |  |  |
| Multiple serious injuries | 747 (5.7%) | 69 (3.5%) |  |  |  |
| AVPU |  |  | 0.19 | 0.14, 0.24 |  |
| Unresponsive | 179 (1.3%) | 7 (0.3%) |  |  |  |
| Pain responsive | 703 (5.3%) | 49 (2.4%) |  |  |  |
| Voice responsive | 257 (1.9%) | 30 (1.5%) |  |  |  |
| Alert | 12,202 (91%) | 1,921 (96%) |  |  |  |
| Dead at 30 days | 704 (5.3%) | 0 (NA%) |  |  |  |
| Dead at 24 hours | 293 (2.2%) | 0 (0%) | 2.2% | 1.9%, 2.5% | <0.001 |
| Dead at 90 days | 757 (6.8%) | 1 (100%) | -93% | -100%, -43% | 0.086 |
| In-hospital mortality | 439 (3.3%) | 0 (0%) | 3.3% | 3.0%, 3.6% | <0.001 |
| Admitted to the hospital | 1,323 (9.9%) | 129 (6.5%) | 3.5% | 2.2%, 4.7% | <0.001 |
| Admitted to the intensive care unit | 58 (0.4%) | 6 (0.3%) | 0.13% | -0.16%, 0.43% | 0.5 |
| Clinician-assigned triage category |  |  | 0.23 | 0.18, 0.27 |  |
| Green | 7,180 (55%) | 1,270 (64%) |  |  |  |
| Yellow | 4,343 (33%) | 459 (23%) |  |  |  |
| Orange | 986 (7.5%) | 166 (8.4%) |  |  |  |
| Red | 559 (4.3%) | 90 (4.5%) |  |  |  |
| *^1^*Median (Q1, Q3); n (%) | | | | | |
| *^2^*Welch Two Sample t-test; Standardized Mean Difference; NA; 2-sample test for equality of proportions with continuity correction | | | | | |
| Abbreviation: CI = Confidence Interval | | | | | |

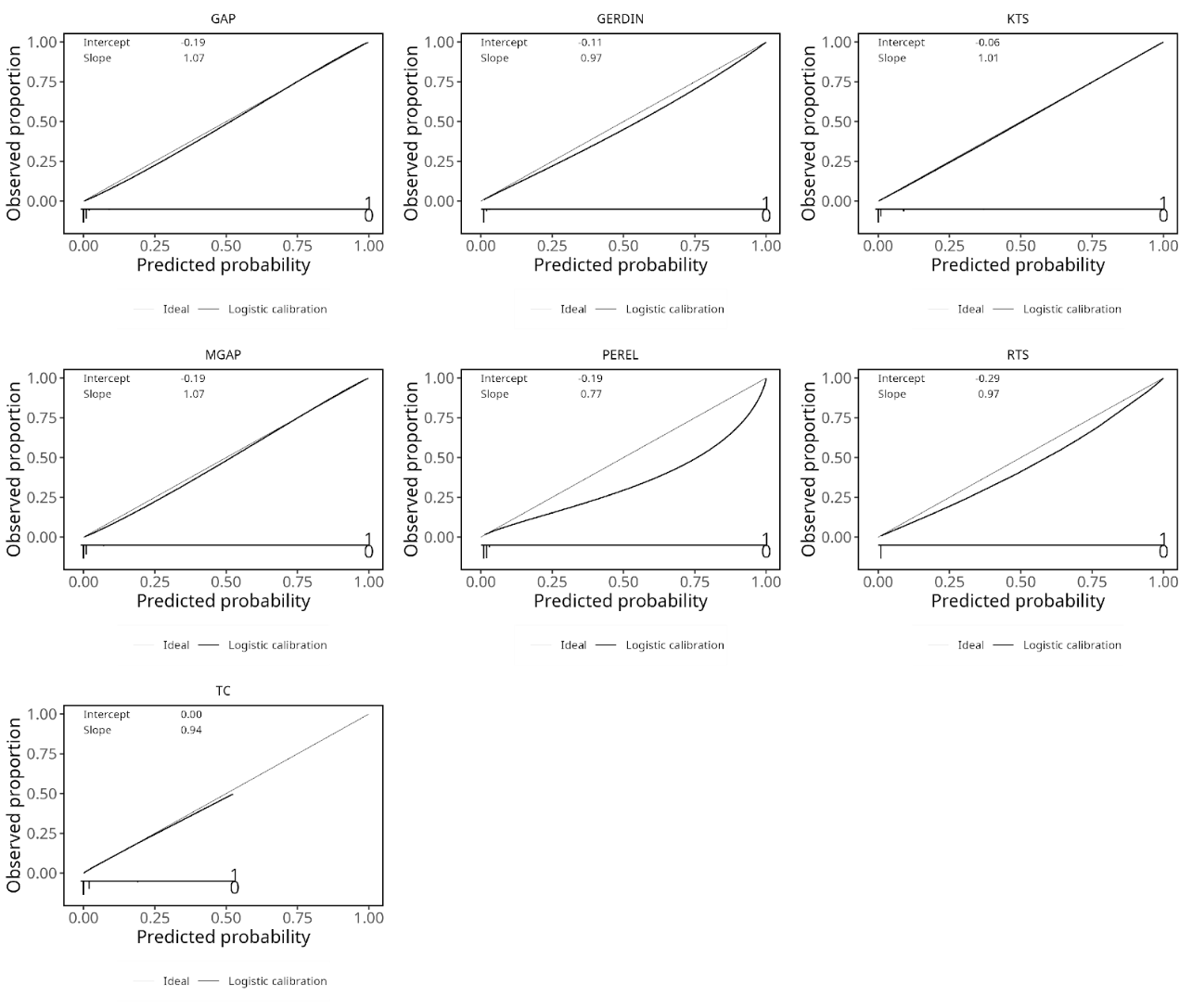

*Appendix 2: Calibration plots for each model*
